# Age, prostate-specific antigen, screening frequency, and metastatic prostate cancer in U.S. Veterans

**DOI:** 10.64898/2026.03.09.26347958

**Authors:** Mehrnaz Siavoshi, Stephen Frochen, Mary Fakunle, Ananta Wadhwa, Ashley-Marie Y. Green-Lott, Anissa V. Bailey, Lorna Kwan, Candace Haroldsen, Atim Effiong, Brent S. Rose, Timothy R. Rebbeck, Hari S. Iyer, Isla P. Garraway

## Abstract

**Importance:** Metastatic prostate cancer (PCa) incidence has increased in U.S. men, partly due to changes in prostate-specific antigen (PSA) screening recommendations. However, few studies have examined contemporary PSA screening practices in large U.S. healthcare systems.

**Objective:** Describe and examine contemporary PSA testing practices associated with metastatic PCa incidence.

**Design:** Cohort study.

**Setting:** Veterans Health Administration.

**Participants:** Veterans diagnosed with prostate needle biopsy (PNBx) between 2015 and 2023 with follow-up through 2024, excluding those with a history of PCa.

**Exposures:** PSA tests were retrieved from the VA corporate data warehouse and categorized by age at first VA PSA (<50, 50-59, ≥60 years) and by longest interval between consecutive VA PSA tests in the 5 years before PNBx (≤24 , >24 months). Clinical, laboratory, pathological, demographic, and Census Block Group-level socioeconomic status data were obtained from the VA Multi-OMICS Analysis Platform for Prostate Cancer (VA-MAPP) database.

**Main Outcomes and Measures:** Multivariable Cox models estimated hazard ratios (HR) from time of first VA PSA to first PNBx, evaluated risk of metastatic (regional or distant) versus localized PCa, or benign diagnosis, adjusted for sociodemographic and clinical covariates. Data were analyzed between July 1, 2023 and November 6, 2025.

**Results:** There were 103,067 participants of whom 20% were <50 years old at first PSA, 31% non-Hispanic Black, 57% non-Hispanic White, and 13% other racial and ethnic groups. Of these, 22% had first PSA value ≤1, 51% had a screening interval ≤24 months, and 4% were diagnosed with metastatic PCa at time of PNBx. Compared to men aged <50 years at first PSA, those 50-59 (aHR 1.08, 95% CI: 1.06-1.11) and ≥60 years (aHR 1.79, 95% CI: 1.74-1.84) had higher metastatic PCa. Men with longer screening intervals had higher metastatic PCa (aHR 1.09, 95% CI: 1.07-1.11). Men aged <50 years with shorter screening intervals had lower metastatic PCa (aHR: 0.10, 95% CI: 0.09-0.12) compared to men aged ≥60 years with longer screening intervals.

**Conclusions and Relevance:** Few male veterans were observed to have the most favorable combinations of age, PSA value, and PSA screening interval in relation to metastatic PCa, suggesting potential for further screening optimization.

## Introduction

Prostate cancer (PCa) is the most frequently diagnosed non-cutaneous cancer and the second leading cause of male cancer-related mortality in the U.S.^1^. Screening for PCa using prostate-specific antigen (PSA) testing is controversial due to conflicting evidence of survival benefits from clinical trials and concerns about harm from overdiagnosis^2–6^. In 2012, the United States Preventative Services Task Force (USPSTF) advised against routine PSA screening for all men^3^. By 2018, as longer-term clinical trial results showed benefits in select populations^7^, guidelines began recommending shared decision-making for PSA screening in men aged 55-69^8^. As PSA testing declined from 2012 to 2018, there was a corresponding reduction in the incidence of localized PCa^9,10^ alongside an increase in metastatic PCa incidence^10,11^ and plateauing of PCa mortality following nearly a decade of decline^12^. These reports have prompted renewed debates regarding how best to screen for PCa in high-risk populations^13–15^.

National PSA screening guidelines focus on age to begin screening, time between screening tests, hereafter referred to as screening interval, and PSA result^8,14^. The National Comprehensive Cancer Institute (NCCN) Prostate Cancer Early Detection and American Urological Association (AUA)/Society of Urologic Oncology (AUA/SUO) early Detection of Prostate Cancer guidelines recommend a baseline PSA test starting at age 45-50 years in average-risk men, and 40-45 years in high-risk men^16,17^. PSA screening every 2-4 years is recommended in average-risk men, and 1-2 years in high-risk men. Lower PSA thresholds can increase overdiagnosis without substantially reducing PCa specific mortality^18,19^. Few studies have evaluated real-world PSA screening patterns in large healthcare systems with respect to these three screening parameters^20,21^.

We sought to describe and examine screening patterns in relation to metastatic PCa at diagnosis within the VA healthcare system. Veterans have higher rates of PCa compared to the general population, due to military occupational exposures or higher access to care^22,23^. However, the VA generally follows USPSTF recommendations with shared decision-making for PSA screening in men ages 55-69 years.

Given these unique risk factors and the potential for more favorable harm/benefit tradeoffs of screening in veteran populations, understanding the relationship between PSA testing patterns and cancer outcomes is a necessary step for optimizing screening recommendations.

## Methods

### Study Design and Population

We conducted a retrospective cohort study using data from the VA Multi-OMICS Analysis Platform for Prostate Cancer (VA-MAPP) database^24^. We captured clinical information from men with at least two VA visits occurring between 2005 to 2020 and who were at risk for PCa, and housed the data in the secure VA Informatics and Computing Infrastructure (VINCI) server (n=11,216,391). Additional clinical, laboratory, pathological, demographic, and Census Block Group-level socioeconomic status data were obtained from the VA Multi-OMICS analysis Platform for Prostate Cancer (VA-MAPP) database as previously described ^22^. We identified each participant’s first VA diagnostic prostate needle biopsy (PNBx) (n=109,110) and restricted the cohort to these first biopsies between 2015 and 2023 (n=103,419). We excluded patients with no VA PSA testing in the 5 years before their first PNBx (n=77) or those with a PCa diagnosis before their first PNBx (n=275), resulting in an analytic sample of 103,067.

The central VA institutional review board and research and development approved the study. We followed the Strengthening the Reporting of Observation Studies in Epidemiology (STROBE) reporting guidelines.

### PSA Test Parameters

PSA tests were retrieved from the “patient lab chem” data table in the VA corporate data warehouse through LOINC codes and key word inclusions (%PSA%, %PROSTAT%, %PROST%SPEC%). Detailed review by coauthors (CH, IPG) were performed to confirm validity of selected key terms to identify PSA tests. Result values were cleaned by excluding erroneous values (e.g. extra decimals) and characters. Only total PSA tests with numerical values were used for analysis based on LOINC 2857-1, 19195-7, and 35741-8.

We evaluated three primary PSA screening parameters: (1) age at first VA PSA test (<50, 50-59, and ≥60 years), (2) value at first VA PSA test (≤1, 1.01-2.50, 2.51-4, and >4 ng/mL), and (3) longest interval between consecutive VA PSA tests in the 5 years prior to biopsy (≤24 months and >24 months). Age was analyzed in categories to better reflect clinical decision making and outcomes for PCa care, which were determined based on thresholds rather than along a continuum. Screening interval length was dichotomized at 24 months because it was most commonly reported in major clinical trials^3,25,26^. For consistency with current recommendations^16,17^, we calculated screening intervals starting at age 45 years for Black veterans and 55 years for all others.

### Outcome

The primary outcome was PCa diagnosis at first PNBx based on ICD-10 codes, categorized as metastatic PCa, localized PCa, or benign (no PCa) diagnosis following biopsy (benign). Metastatic PCa was determined using the metastasis flag in the Prostate Cancer Data Core in VINCI, defined as evidence of metastasis to lymph nodes or distant spread to other organs as described previously.^27^ Survival time was calculated in years from the first VA PSA test to first VA PNBx.

### Covariates

Covariates included race/ethnicity (non-Hispanic White, non-Hispanic Black, Other), Agent Orange exposure based on VA-determined probable exposure^22^, service-connected disability percentage (0-24.9, 25-49.9, 50-74.9, 75-100), Body Mass Index (BMI) at PNBx (obese, non-obese), and the Census Block Group Area Deprivation Index (ADI) State Decile at PNBx (1-2, 3-4, 5-6, 7-8, 9-10)^28^, as a measure of neighborhood socioeconomic disadvantage. We considered Charlson Comorbidity Index (CCI) as a covariate but omitted it from main analyses because it had no impact on results.

### Statistical Analyses

Cohort characteristics were summarized across PCa outcomes. Kaplan-Meier survival curves and log-rank tests were calculated to compare the time from first VA PSA test to metastatic PCa at first VA PNBx stratified by levels of each screening parameter. For our primary analysis, we fit covariate-adjusted Cox proportional hazards models to estimate associations between PSA screening parameters and metastatic PCa at diagnosis. We first assessed associations of each PSA screening parameter separately and then with mutual adjustment for all PSA screening factors. The proportional hazards assumption was evaluated using Schoenfeld residuals^29^, which confirmed no significant violations.

For our secondary analysis to identify combinations of screening factors associated with lowest metastatic PCa rates, we fit Poisson regression models using the same covariates as in the Cox models, with robust variance and an offset term for log person-time to estimate absolute rates of metastatic PCa diagnosis^30^. Poisson models were used because they provided rate estimates on the absolute scale instead of the relative scale. In each Poisson model, the highest risk group was selected as the reference to facilitate clinical interpretation. Three separate interaction models were fitted: (1) age at first PSA by first PSA value (joint reference: age ≥60 and PSA >4 ng/mL); (2) PSA testing interval by first PSA value (joint reference: interval >24 months and PSA >4 ng/mL); and (3) age at first PSA by PSA testing interval (joint reference: age ≥60 and interval >24 months). Relative rates and 95% confidence intervals were estimated for each composite exposure category compared to the joint reference group. Heatmaps were generated for each modified Poisson interaction model to illustrate the joint effects of exposure combinations compared to the respective reference group.

### Sensitivity Analysis

By selecting our cohort based on PNBx, which occurred after PSA screening started, we may have induced selection bias^24^. Under this scenario, we would expect the association between screening parameters and PCa incidence to be biased, leading to an overestimated hazard ratio or rate ratio. We calculated stabilized inverse probability of censoring weights (IPCW)^31^ for all 11,216,391 patients in VA-MAPP, separately for age at first PSA and first PSA value. IPCW for screening interval could not be calculated for those without a biopsy because of the requirement of a 5-year lookback. IPCW were estimated from logistic regression models to predict biopsy conditional on race, ADI, BMI at first PSA, service-connected percent, and Agent Orange exposure, plus either PSA screening parameter. The numerator was estimated from the logistic regression models to predict the probability of biopsy conditional only on the exposure of interest. Stabilized weights were truncated at the 1^st^ and 99^th^ percentiles to improve stability, and covariate balance was assessed using standardized mean differences which showed good balance^32^. We then refit separate Cox proportional hazards models for each PSA screening parameter using the truncated, exposure-specific weights to estimate hazard ratios in a pseudopopulation adjusted for selection into the biopsy cohort^32^.

All statistical analyses were conducted using SAS Enterprise Guide 8.3 (SAS Institute Inc., Cary, NC). Statistical significance was set at α = 0.05.

## Results

### Cohort Characteristics

The analytic cohort included 103,067 veterans who underwent their first VA PNBx between 2015 and 2023. Of these, 3,773 (3.7%) were diagnosed with metastatic PCa, 46,132 (44.8%) were diagnosed with localized PCa, and 53,162 (51.6%) had benign biopsies (Table 1). Veterans diagnosed with metastatic PCa were older at the time of first PSA testing, with 46.4% ≥60 years at first PSA testing compared to 36.4% of localized cases and 32.6% of benign cases (p < 0.001; Table 1). The cohort was racially diverse, with non-Hispanic White veterans comprising the majority across all diagnostic groups (57.6% of metastatic cases, 54.9% of localized cases, and 57.8% of benign cases). Veterans diagnosed with metastatic and localized PCa had higher prevalence of severe service-connected disability (75-100%): 35.8% and 37.3%, respectively, versus 31.1% in the benign group (p < 0.001; Table 1). Median follow-up time was 18 years (IQR 12-23) overall, with limited variation by metastatic status at time of PNBx.

**Table 1:** Cohort characteristics and PSA testing factors by diagnosis at first VA prostate needle biopsy (N = 103,067). Abbreviations: ADI=Area Deprivation Index, ADT=Androgen Deprivation Therapy, CCI=Charlson Comorbidity Index, PCa=Prostate Cancer, PNBx=Prostate Needle Biopsy, PSA=Prostate-specific Antigen, IQR=Interquartile Range, ^a^Chi-Square p-value; ^b^Kruskal-Wallis p-value

|  | Total<br>(N = 103,067) | Diagnosis at First VA PNBx |  |  | p-value |
| --- | --- | --- | --- | --- | --- |
|  |  | Metastatic<br>(n = 3,773) | Localized<br>(n = 46,132) | Benign<br>(n = 53,162) |  |
| **Race/Ethnicity, n (%)** |  |  |  |  | <0.001<sup>a</sup> |
| Non-Hispanic Black | 31546 (30.6) | 1183 (31.4) | 15553 (33.7) | 14810 (27.9) |  |
| Non-Hispanic White | 58264 (56.5) | 2173 (57.6) | 25368 (54.9) | 30723 (57.8) |  |
| Other | 13277 (12.9) | 417 (11.1) | 5204 (11.3) | 7629 (14.4) |  |
| **Agent Orange Exposure, n (%)** | 18405 (17.9) | 704 (18.7) | 9046 (19.6) | 8655 (16.3) | <0.001<sup>a</sup> |
| **Service-Connected Percent, n (%)** |  |  |  |  | <0.001<sup>a</sup> |
| 0-24.9 | 49305 (47.8) | 1961 (52.0) | 21084 (45.7) | 26260 (49.4) |  |
| 25-49.9 | 6059 (5.9) | 168 (4.5) | 2546 (5.5) | 3345 (6.3) |  |
| 50-74.9 | 12604 (12.2) | 293 (7.8) | 5313 (11.5) | 6998 (13.2) |  |
| 75-100 | 35099 (34.1) | 1351 (35.8) | 17189 (37.3) | 16559 (31.1) |  |
| **Deaths over follow-up, n (%)** | 13344 (12.9) | 1660 (44.0) | 6805 (14.8) | 4879 (9.2) | <0.001<sup>a</sup> |
| **BMI at First PNBx, mean (SD)** | 29.6 (5.64) | 29.1 (5.81) | 29.8 (5.79) | 29.5 (5.50) | <0.001<sup>b</sup> |
| **BMI at First PNBx, n (%)** |  |  |  |  | <0.001<sup>a</sup> |
| Non-Obese | 56114 (57.2) | 2243 (60.4) | 25271 (56.2) | 28699 (57.9) |  |
| Obese | 41970 (42.8) | 1475 (39.7) | 19646 (43.7) | 20849 (42.2) |  |
| Missing | 4983 | 55 | 1215 | 3713 |  |
| **CCI at First PSA, n (%)** |  |  |  |  | 0.027<sup>a</sup> |
| 0 | 96145 (93.3) | 3537 (93.7) | 43014 (93.2) | 49594 (93.3) |  |
| 1-2 | 5915 (5.7) | 200 (5.3) | 2621 (5.7) | 3094 (5.8) |  |
| 3+ | 1007 (1.0) | 36 (1.0) | 497 (1.1) | 474 (0.9) |  |
| **State ADI at First PNBx, n (%)** |  |  |  |  | <0.001<sup>a</sup> |
| 0-2 | 10143 (9.8) | 401 (10.6) | 4778 (10.4) | 4964 (9.3) |  |
| 3-4 | 15243 (14.8) | 545 (14.4) | 7279 (15.8) | 7419 (14.0) |  |
| 5-6 | 16851 (16.3) | 641 (17.0) | 8136 (17.6) | 8074 (15.2) |  |
| 7-8 | 17287 (16.8) | 704 (18.7) | 8430 (18.3) | 8153 (15.3) |  |
| 9-10 | 17170 (16.7) | 722 (19.1) | 8491 (18.4) | 7957 (15.0) |  |
| Null | 26373 (25.6) | 760 (20.1) | 9018 (19.5) | 16595 (31.2) |  |
| **PCa Treatment, n (%)** | 13880 (26.2) | 858 (22.7) | 12138 (26.3) | 884 (28.8) | <0.001<sup>a</sup> |

**Table 1: Cohort characteristics and PSA testing factors by diagnosis at first VA prostate needle biopsy (N = 103,067).**
|  |  |  |  |  |  |
| --- | --- | --- | --- | --- | --- |
| Prostatectomy | 11267 (21.3) | 806 (21.4) | 9899 (21.5) | 562 (18.3) |  |
| Radiation | 10153 (19.2) | 1926 (51.0) | 7739 (16.8) | 488 (15.9) |  |
| ADT Only | 17673 (33.4) | 183 (4.9) | 16356 (35.5) | 1134 (37.0) |  |
| Other/Unknown | 50094 | 0 | 0 | 50094 |  |
| Never Diagnosed |  |  |  |  |  |
| **Years of Follow Up**, median (IQR) | 18 (12, 23) | 17 (12, 22) | 18 (13, 23) | 17 (12, 23) | <0.001<sup>b</sup> |
| **PSA Screening Parameters** |  |  |  |  |  |
| **Age at First PSA**, years, n (%) |  |  |  |  | <0.001<sup>a</sup> |
| <50 | 20233 (19.6) | 456 (12.1) | 7999 (17.3) | 11778 (22.2) |  |
| 50-59 | 46958 (45.6) | 1567 (41.5) | 21335 (46.2) | 24056 (45.3) |  |
| ≥60 | 35876 (34.8) | 1750 (46.4) | 16798 (36.4) | 17328 (32.6) |  |
| **First PSA Value**, ng/mL, n (%) |  |  |  |  | <0.001<sup>a</sup> |
| ≤1 | 22190 (21.5) | 1137 (30.1) | 10651 (23.1) | 10402 (19.6) |  |
| 1.01-2.50 | 42722 (41.5) | 1271 (33.7) | 19486 (42.2) | 21965 (41.3) |  |
| 2.51-4 | 17713 (17.2) | 404 (10.7) | 7378 (16.0) | 9931 (18.7) |  |
| >4 | 20442 (19.8) | 961 (25.5) | 8617 (18.7) | 10864 (20.4) |  |
| **PSA Testing Interval**, n (%) |  |  |  |  | <0.001<sup>a</sup> |
| Gap ≤24 months | 52939 (51.4) | 1210 (32.1) | 23492 (50.9) | 28237 (53.1) |  |
| Gap >24 months | 50128 (48.6) | 2563 (67.9) | 22640 (49.1) | 24925 (46.9) |  |

### Associations of Screening Parameters with Metastatic PCa

Kaplan-Meier curves demonstrated significant differences in the probability of metastatic PCa at PNBx across all three screening parameters (log-rank p < 0.001 for each comparison; Figure 1, Table 2). Veterans aged ≥60 years at first PSA had higher incidence of metastatic PCa over follow-up (766.8 cases per 100,000 person-years) compared to ages 50-59 years (289.8 cases) and <50 years (164.1) (Table 2, Figure 1A). The cumulative incidence of metastatic PCa in this cohort was highest among those with >4 ng/mL (1,420.9 cases per 100,000 person-years), compared to those with ≤1 ng/mL (346.1), 1-2.5 ng/mL (246.6) and 2.5-4 ng/mL (298.7) (Figure 1B). Veterans with PSA testing intervals >24 months had higher metastatic PCa over follow-up compared to those testing intervals ≤24 months (582.3 v 199.4 cases per 100,000 person-years) (Figure 1C).

**Figure 1:**
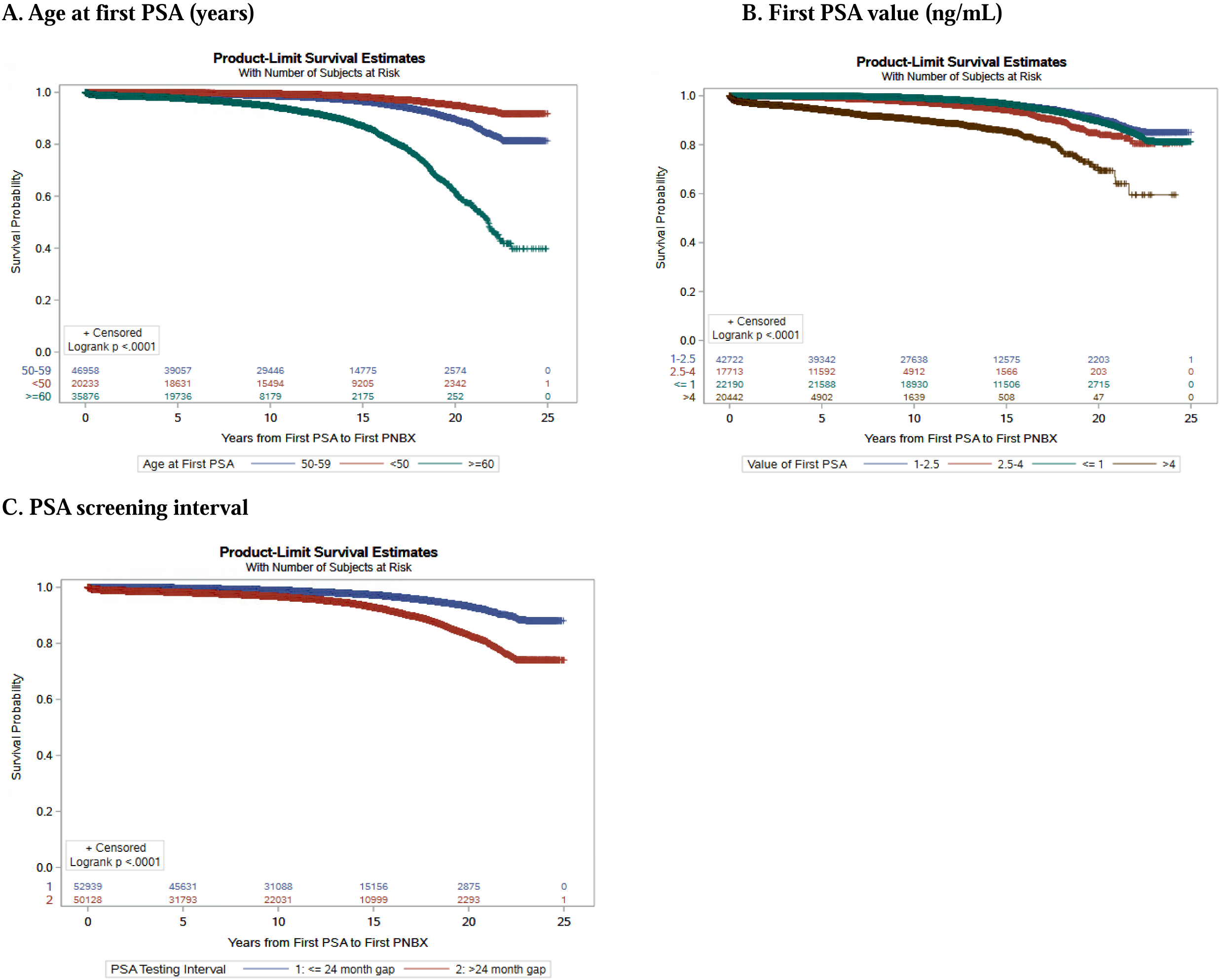
Kaplan-Meier curves for time from first PSA to diagnosis of metastatic prostate cancer at time of biopsy stratified by screening factors. Abbreviations: PNBx=Prostate Needle Biopsy, PSA=Prostate-specific Antigen. Log-rank p-values <0.001 for all co parisons

**Table 2.** Cumulative incidence of metastatic prostate cancer and median time to prostate needle biopsy by screening factors. Abbreviations: PCa=Prostate Cancer, PSA=Prostate-specific Antigen, IQR=Interquartile Range, All Chi-square tests for cumulative incidence p < 0.001

| **PSA Screening Factor** | **Cases of metastatic PCa** | **Population at Risk** | **Cumulative incidence of metastatic PCa (%)** | **Person-years** | **Incidence rate per 100,000** |
| --- | --- | --- | --- | --- | --- |
| **Age at First PSA (years)** |  |  |  |  |  |
| <50 | 456 | 20,233 | 2.3 | 277,821 | 164.1 |
| 50-59 | 1,567 | 46,958 | 3.3 | 540,808 | 289.8 |
| ≥60 | 1,750 | 35,876 | 4.9 | 228,208 | 766.8 |
| **First PSA Value (ng/mL)** |  |  |  |  |  |
| ≤1 | 1,137 | 22,190 | 5.1 | 328,514 | 346.1 |
| 1.01-2.50 | 1,271 | 42,722 | 3.0 | 515,418 | 246.6 |
| 2.51-4 | 404 | 17,713 | 2.3 | 135,273 | 298.7 |
| >4 | 961 | 20,442 | 4.7 | 67,632 | 1420.9 |
| **PSA Testing Interval (months)** |  |  |  |  |  |
| ≤24 | 1,210 | 52,939 | 2.3 | 606,706 | 199.4 |
| >24 | 2,563 | 50,128 | 5.1 | 440,131 | 582.3 |

In the models adjusting for each screening parameter individually, all three factors showed strong associations with metastatic PCa diagnosis (Supplementary Table 1). First PSA value >4 ng/mL versus ≤1 ng/mL had the strongest unadjusted association (aHR 8.53, 95% CI: 8.35-8.72), followed by age ≥60 years versus <50 at first PSA (aHR 3.55, 95% CI: 3.48-3.62), while testing interval of >24 months versus ≤24 months showed a more modest effect (aHR 1.25, 95% CI 1.24-1.27).

When all three screening parameters were included simultaneously, associations were attenuated but remained significant (Figure 2, Supplementary Table 2). Compared to age <50 years at first PSA, veterans aged 50-59 (aHR 1.08, 95% CI 1.06-1.11) and ≥60 years (aHR 1.79, 95% CI 1.74-1.84) had higher rate of metastatic PCa at PNBx. Compared to men with PSA values ≤1 ng/mL, those with 1.01-2.50 ng/mL (aHR 1.63, 95% CI: 1.59-1.67), 2.51-4 ng/mL (aHR 3.46, 95% CI 3.36-3.57), and >4 ng/mL (aHR 8.11, 95% CI: 7.86-8.36) had higher rates of metastatic PCa risk. Compared to men with a PSA testing interval ≤24 months, those with intervals >24 months had higher rate of metastatic PCa diagnosis (aHR 1.09, 95% CI: 1.07-1.11).

**Figure 2:**
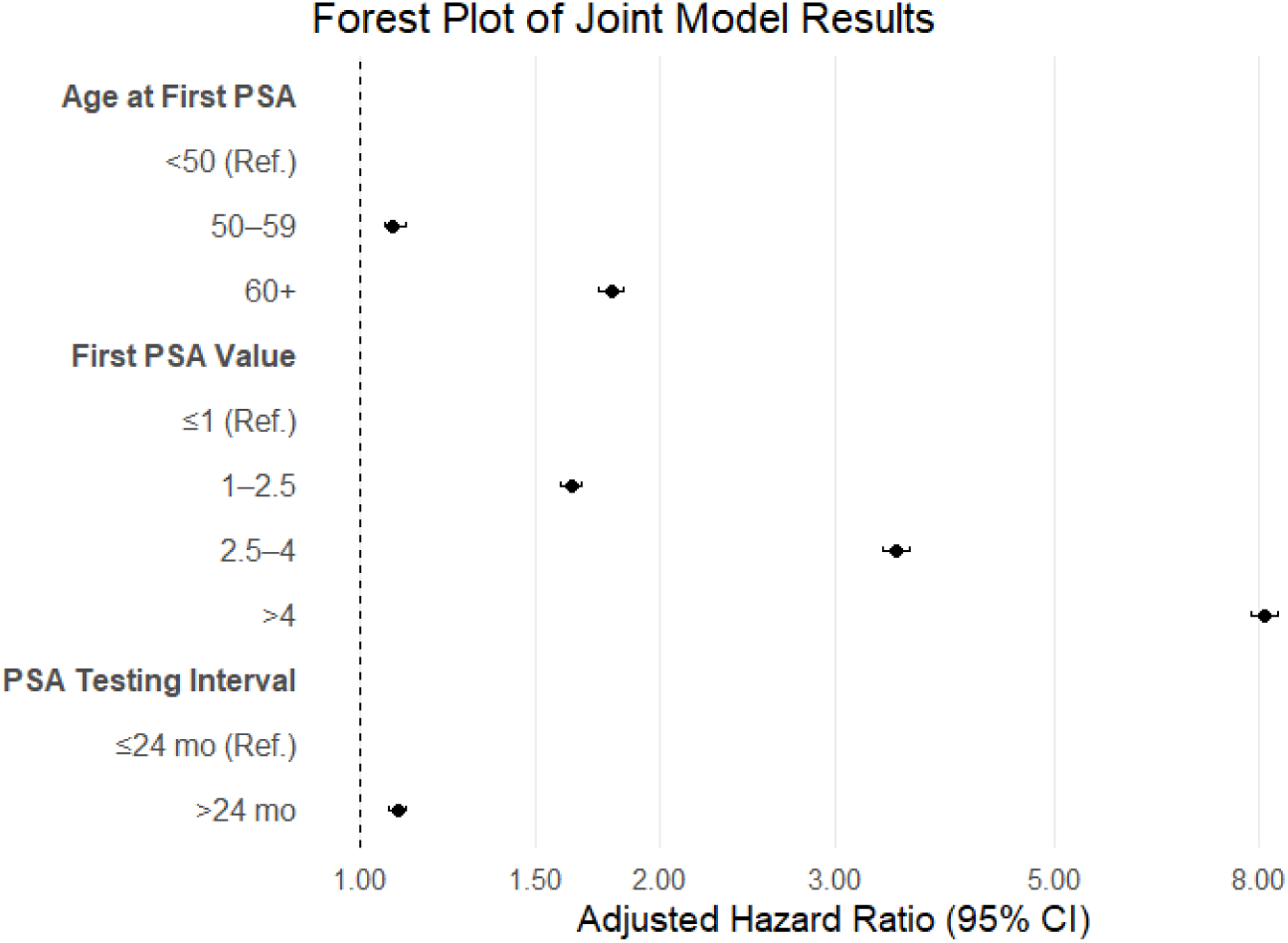
Adjusted hazard ratios for associations of Age at First PSA, First PSA value, and PSA Testing Interval with metastatic prostate cancer in men seeking care in the Veteran Administration. Abbreviations: PNBx=Prostate Needle Biopsy, PSA=Prostate-specific Antigen. Points represent point estimate, lines represent 95% confidence intervals. Results from multivariable Cox models adjusted for Age at first PSA, First PSA value, PSA testing interval, Agent Orange exposure, Service-Connected percent, State ADI at first PNBx, Race (Non-Hispanic Black vs Other), and BMI (obese vs not obese) at first PNBx.

### Interactions Between Screening Parameters and Metastatic PCa

Multivariable Poisson regression models with interaction terms revealed complex risk stratification patterns (Figure 3, Supplementary Tables 3-5). For the interaction between age at first PSA and first PSA value, first PSA value was the primary driver of risk variation as demonstrated by the gradient differences across PSA categories within each age group (Figure 3A). The lowest risk occurred in veterans aged <50 years with PSA 1.01-2.50 ng/mL (aRR: 0.07, 95% CI: 0.06-0.09). All combinations of age ≤59 years and PSA ≤4 ng/mL had substantially lower metastatic PCa risk than the joint reference group (age ≥60 years and PSA >4 ng/mL).

**Figure 3:**
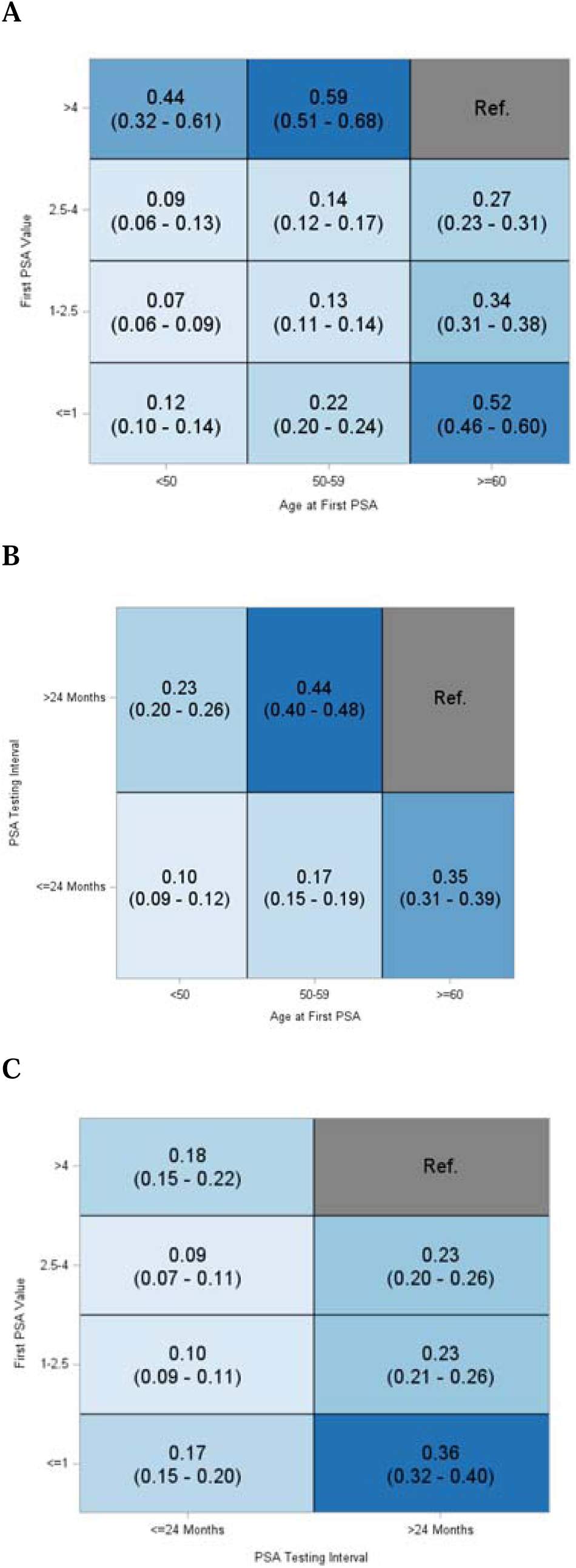
Relative Rates for Interactions between First PSA Value, Age at First PSA, and PSA testing interval in men receiving care in the Veterans Health Administration. Abbreviations: PSA=Prostate-specific Antigen. Legend: Relative rates calculated from multivariable Poisson regression models with an interaction between First PSA value (ng/mL), Age at First PSA (years), and PSA testing Interval (months), adjusting for the third variable. All Poisson models were adjusted for Agent Orange exposure, Service-Connected percent, State ADI at first PNBX, Race (Non-Hispanic Black vs Other), and BMI (obese vs not obese) at first PNBx.

For the interaction between age at first VA PSA and testing interval, risk increased progressively with both older age and longer testing gaps (Figure 3B). The lowest risk occurred among veterans aged <50 years with an interval ≤24 months (aRR: 0.10, 95% CI: 0.09-0.12) relative to the joint reference group (age ≥60 years and gap >24 months).

For the interaction between first VA PSA value and testing interval, PSA testing interval showed the strongest effect with differences across interval categories within each PSA value group (Figure 3C). The lowest risk occurred among veterans with a testing interval ≤ 24 months and PSA of 2.50-4 ng/mL (aRR: 0.09, 95% CI: 0.07-0.11) as compared to the joint reference group (gap >24 months and PSA >4 ng/mL). Interestingly, among those with gaps >24 months, the lowest PSA category (≤1 ng/mL, aRR 0.17, 95% CI: 0.15-0.20) had relatively higher risk than those with PSA 1.01-2.50 ng/mL (aRR: 0.09, 95% CI: 0.08-0.11) possibly reflecting delayed diagnosis in men with initially low PSA who were screened infrequently.

### Sensitivity Analysis

In IPCW analyses to assess robustness of results to potential selection bias, the effective sample sizes were 47.7% of the original cohort for age at first PSA and 72.2% for first PSA value. The weighted analysis yielded results consistent with the primary findings, with slight attenuation of the effect estimates for age at first PSA and slight elevation for first PSA value (Supplementary Table 6).

## Discussion

In this cohort study of men undergoing diagnostic PNBx within the VA healthcare system, older age at first VA PSA test, longer testing intervals, and higher initial PSA values were independently associated with metastatic disease at PCa diagnosis. The lowest metastatic PCa risk was observed in veterans aged <50 years at first VA PSA test and initial PSA value of 1.01-2.50 ng/mL. Veterans aged <50 with testing interval ≤24 months also demonstrated very low risk, as did those with PSA 2.51-4 ng/mL and interval ≤24 months.

Understanding how combinations of PSA screening parameters (age, interval, value) interact in relation to risks of metastatic PCa is essential for balancing harm-benefit tradeoffs^14,33–37^. In response to lower PSA screening rates in the years following USPSTF’s guideline changes in 2012^38^, VA studies have reported increased metastatic PCa risk^39^. Previous studies have also found that more frequent PSA screening (shorter intervals between tests) was associated with lower risk of metastatic PCa, consistent with our findings^33^, but at the cost of unnecessary biopsies and overtreatment^40^. We found that successive PSA tests >24 months apart were associated with a nearly 10% increased risk of metastatic PCa, which was exacerbated with delayed age at initial screening. Veterans with longer screening intervals and who started screening at older ages were at highest risk of metastatic presentation, emphasizing the importance of not only regular but earlier testing in detecting and reducing metastatic PCa at diagnosis. Further, when factoring in PSA values, the risk of metastatic PCa increased for each progressively higher categorical PSA level in veterans with consecutive tests, regardless of interval, with the greatest risk of metastatic PCa occurring in patients with longer intervals between VA tests and initial PSA values >4 ng/mL.

Disparities associated with screening factors are often accentuated among veterans, who experience elevated baseline metastatic PCa risk, particularly among non-Hispanic Black veterans and veterans with service-related exposures, such as Agent Orange, which predicts higher PCa risk in this population^22,41,42^. Polygenic risk scores may offer opportunities to further tailor screening based on individual risk, and inform shared decision-making conversations between providers and patients^42–45^. Enhanced risk-stratified monitoring in veterans could increase PCa incidence even while reducing aggressive disease^42^. Black men and individuals residing in more deprived neighborhoods are at greater risk of metastatic PCa in non-VA healthcare settings^27,46,47^. Because VA guidelines are adopted centrally and applied throughout the system, changes in screening practice may be more consistently applied and therefore lead to greater impacts within this healthcare system. Our analyses can therefore serve as a starting point for modeling impacts of different screening strategies that can estimate the harm-benefit tradeoffs in relation to treatment side effects and survival in diverse populations^48,49^.

Our study had some limitations. Our data did not include PSA tests that occurred outside of the VA, which could have introduced misclassification bias if missing tests are correlated with metastatic PCa incidence. Restricting to men with PNBx could have also introduced selection bias if more aggressive testing by clinicians and medical providers, as well as access to different forms of healthcare, was correlated with biopsy and subsequent metastatic PCa. Our sensitivity analyses using IPCW yielded similar results to our main analyses, suggesting that selection bias alone was insufficient to explain our findings. Our sample may have overrepresented certain groups within the VA subpopulation, including veterans who used and/or were more engaged in VA care, those with higher service-connected disability ratings who receive priority access to care at VA, as well as urban VA patients who typically have greater access to primary and specialty care at VA. Further, VA has historically had a greater proportion of Black patients compared to the general population, who may undergo biopsy with greater frequency at VA as patients with higher baseline PCa risk^50^. This may limit the generalizability of our findings to the broader U.S. population. Lastly, unmeasured clinical or provider-related factors may have contributed to confounding bias in our results. Study strengths included a large and diverse cohort of veterans over an extended follow-up period, utilizing real-world administrative data from a nationally integrated U.S. healthcare system.

In summary, our study provides a contemporary look at relationships between age at first PSA, PSA screening interval, and PSA value at first test in a large U.S. population of men at risk of PCa. Our findings support continued efforts to focus screening in high-risk men to avoid harms of overtreatment, and to tailor strategies of age, screening interval and PSA value to optimize screening in different groups of men. These results can inform modeling studies to evaluate potential benefits of optimized screening strategies based on genetics and other prognostic factors.

## Author contributions

Conceptualization: MS, HSI, IPG; Funding acquisition: IPG; Data curation: CH, AE; Formal analysis: MS, SF; Writing – original draft: MS, SF; Writing – review and editing: all authors

## Data Availability

All data produced in the present study are available upon reasonable request to the authors.

