## Supplemental Material for "Age, prostate-specific antigen, screening frequency, and metastatic prostate cancer in U.S. Veterans"

S**upplementary Table 1: Adjusted hazard ratios for separate associations of Age at First PSA, First PSA Value, and PSA Testing Interval with Metastatic Prostate Cancer**

**Supplementary Table 2: Adjusted hazard ratios for associations of Age at First PSA, First PSA value, and PSA Testing Interval with metastatic prostate cancer at time of prostate needle biopsy in men seeking care in the Veterans Administration**

**Supplementary Table 3: Relative Rates for Interaction between First PSA Value and Age at First PSA**

**Supplementary Table 4: Relative Rates for Interaction between Age at First PSA and PSA Testing Interval**

**Supplementary Table 5: Relative Rates for Interaction between First PSA Value and PSA Testing Interval**

**Supplementary Table 6: Sensitivity Analysis Comparison of Unweighted and Inverse Probability of Censoring Weighted Cox Model to Adjust for Selection Bias**

S**upplementary Table 1: Adjusted hazard ratios for separate associations of Age at First PSA, First PSA Value, and PSA Testing Interval with Metastatic Prostate Cancer**

|  | **Model 1 Age at First PSA**  **HR (95% CI)** | **Model 2 First PSA Value**  **HR (95% CI)** | **Model 3 PSA Testing Interval**  **HR (95% CI)** |
| --- | --- | --- | --- |
| **Age at First PSA (years)**  <50  50-59  ≥60 | Ref.  1.40 (1.37 - 1.42)  3.55 (3.48 - 3.62) |  |  |
| **First PSA Value (ng/mL)**  ≤1  1.01-2.50  2.51-4  >4 |  | Ref.  1.54 (1.52 – 1.57)  3.31 (3.24 – 3.39)  8.53 (8.35 – 8.72) |  |
| **PSA Testing Interval**  Gap ≤24 months  Gap >24 months |  |  | Ref.  1.25 (1.24 – 1.27) |
| **Covariates** | | | |
| **Agent Orange Exposure**  No  Yes | Ref.  0.91 (0.89 - 0.92) | Ref.  1.06 (1.04 – 1.08) | Ref.  1.17 (1.15 – 1.19) |
| **Service-Connected Percent**  0-24.9%  25-49.9%  50-74.9%  75-100% | Ref.  1.00 (0.97 - 1.03)  0.94 (0.92 - 0.96)  0.88 (0.87 - 0.89) | Ref.  0.97 (0.95 – 1.00)  0.90 (0.88 – 0.92)  0.86 (0.85 – 0.87) | Ref.  0.91 (0.89 – 0.94)  0.83 (0.81 – 0.85)  0.77 (0.76 – 0.78) |
| **State ADI at First PNBx**  0-2  3-4  5-6  7-8  9-10  Null | Ref.  1.01 (0.98 - 1.03)  0.96 (0.93 - 0.98)  0.93 (0.91 - 0.96)  0.91 (0.89 - 0.94)  0.76 (0.75 - 0.78) | Ref.  1.01 (0.99 – 1.04)  0.96 (0.93 – 0.98)  0.93 (0.90 – 0.95)  0.89 (0.87 – 0.92)  0.76 (0.74 – 0.78) | Ref.  0.99 (0.96 – 1.01)  0.93 (0.91 – 0.95)  0.89 (0.87 – 0.92)  0.97 (0.94 – 0.89)  0.73 (0.71 – 0.75) |
| **Race**  Non-Hispanic Black  Other | Ref.  0.96 (0.94 - 0.97) | Ref.  1.03 (1.02 – 1.05) | Ref.  1.15 (1.13 – 1.16) |
| **BMI at PNBx**  Not Obese  Obese | Ref.  1.18 (1.16 - 1.19) | Ref.  1.15 (1.14 – 1.17) | Ref.  1.11 (1.10 – 1.13) |

Abbreviations: PNBx=Prostate Needle Biopsy, PSA=Prostate-specific Antigen. Legend: Hazard ratios calculated from multivariable Cox regression models with for First PSA value (ng/mL), PSA testing Interval (months), Age at First PSA (years) with metastatic PCa and follow-up through PNBx. All Cox models were adjusted for Agent Orange exposure, service-connected percent, State ADI at first PNBx, race (Non-Hispanic Black vs other), and BMI (obese vs not obese) at first PNBx. There were 4,983 observations dropped due to missing BMI observations.

**Supplementary Table 2: Adjusted hazard ratios for associations of Age at First PSA, First PSA value, and PSA Testing Interval with metastatic prostate cancer at time of prostate needle biopsy in men seeking care in the Veterans Administration**

|  | **Single Variable Model  aHR (95% CI)** | **Mutually Adjusted Model aHR (95% CI)** |
| --- | --- | --- |
| **Age at First PSA (years)**  <50  50-59  ≥60 | Ref.  1.40 (1.37 - 1.42)  3.55 (3.48 - 3.62) | Ref.  1.08 (1.06 – 1.11)  1.79 (1.74 – 1.84) |
| **First PSA Value (ng/mL)**  ≤1  1.01-2.50  2.51-4  >4 | Ref.  1.54 (1.52 – 1.57)  3.31 (3.24 – 3.39)  8.53 (8.35 – 8.72) | Ref.  1.63 (1.59 – 1.67)  3.46 (3.36 – 3.57)  8.11 (7.86 – 8.36) |
| **PSA Testing Interval**  Gap ≤24 months  Gap >24 months | Ref.  1.25 (1.24 – 1.27) | Ref.  1.09 (1.07 – 1.11) |

Abbreviations: PNBx=Prostate Needle Biopsy, PSA=Prostate-specific Antigen. Legend: Hazard ratios calculated from multivariable Cox regression models with for First PSA value (ng/mL), PSA testing Interval (months), Age at first PSA (years) with metastatic PCa and follow-up through PNBx. All Cox models were adjusted for Agent Orange exposure, service-connected percent, State ADI at first PNBx, race (non-Hispanic Black vs other), and BMI (obese vs not obese) at first PNBx.

**Supplementary Table 3: Relative Rates for Interaction between First PSA Value and Age at First PSA**

|  | **aRR (95% CI)** |
| --- | --- |
| **Interaction Terms** | |
| Age <50 years  PSA ≤1 ng/mL  PSA 1.01-2.50 ng/mL  PSA 2.51-4 ng/mL  PSA >4 ng/mL | 0.12 (0.10 – 0.14)  0.07 (0.06 – 0.09)  0.09 (0.06 – 0.13)  0.44 (0.31 – 0.62) |
| Age 50-59 years  PSA ≤1 ng/mL  PSA 1.01-2.50 ng/mL  PSA 2.51-4 ng/mL  PSA >4 ng/mL | 0.22 (0.20 – 0.24)  0.13 (0.11 – 0.14)  0.14 (0.12 – 0.17)  0.53 (0.45 – 0.64) |
| Age ≥60 years  PSA ≤1 ng/mL  PSA 1.01-2.50 ng/mL  PSA 2.51-4 ng/mL  PSA >4 ng/mL (highest risk group) | 0.52 (0.45 – 0.61)  0.34 (0.31 – 0.38)  0.27 (0.23 – 0.31)  Ref |
| **Other Screening Factor** | |
| PSA Testing Interval  Gap ≤24 months  Gap >24 months | Ref  2.65 (2.48 – 2.84) |

Abbreviations: aRR=adjusted relative rate, PSA=Prostate-specific Antigen. Legend: Relative rates calculated from multivariable Poisson regression models with an interaction between Age at First PSA (years) and First PSA value (ng/mL), adjusting for PSA testing interval. All Poisson models were adjusted for Agent Orange exposure, service-connected percent, State ADI at first PNBx, race (non-Hispanic Black vs other), and BMI (obese vs not obese) at first PNBx.

**Supplementary Table 4: Relative Rates for Interaction between Age at First PSA and PSA Testing Interval**

|  | **aRR (95% CI)** |
| --- | --- |
| **Interaction Terms** | |
| Age <50 years  Gap ≤24 months  Gap >24 months | 0.10 (0.09 – 0.12)  0.23 (0.20 – 0.26) |
| Age 50-59 years  Gap ≤24 months  Gap >24 months | 0.17 (0.15 – 0.19)  0.44 (0.40 – 0.47) |
| Age ≥60 years  Gap ≤24 months  Gap >24 months (highest risk group) | 0.35 (0.32 – 0.39)  Ref |
| **Other Screening Factor** | |
| First PSA Value  ≤1 ng/mL  1.01-2.50 ng/mL  2.51-4 ng/mL  >4 ng/mL | Ref  0.62 (0.57 – 0.67)  0.59 (0.52 – 0.66)  2.20 (2.01 – 2.42) |

Abbreviations: aRR=adjusted relative rate, PSA=Prostate-specific Antigen. Legend: Relative rates calculated from multivariable Poisson regression models with an interaction between Age at First PSA (years) and PSA testing Interval (months) adjusting for First PSA value (ng/mL). All Poisson models were adjusted for Agent Orange exposure, service-connected percent, State ADI at first PNBx, race (non-Hispanic Black vs other), and BMI (obese vs not obese) at first PNBx.

**Supplementary Table 5: Relative Rates for Interaction between First PSA Value and PSA Testing Interval**

|  | **aRR (95% CI)** |
| --- | --- |
| **Interaction Terms** | |
| PSA ≤1 ng/mL  Gap ≤24 months  Gap >24 months | 0.17 (0.15 – 0.20)  0.10 (0.09 – 0.11) |
| PSA 1.01-2.50 ng/mL  Gap ≤24 months  Gap >24 months | 0.09 (0.08 – 0.11)  0.18 (0.15 – 0.22) |
| PSA 2.50-4 ng/mL  Gap ≤24 months  Gap >24 months | 0.36 (0.32 – 0.40)  0.23 (0.21 – 0.26) |
| PSA >4 ng/mL  Gap ≤24 months  Gap >24 months | 0.23 (0.20 – 0.26)  Ref |
| **Other Screening Factor** | |
| Age at First PSA  <50 years  50-59 years  ≥60 years | Ref  1.79 (1.61 – 1.99)  4.01 (3.57 – 4.50) |

Abbreviations: aRR=adjusted relative rate, PSA=Prostate-specific Antigen. Legend: Relative rates calculated from multivariable Poisson regression models with an interaction between First PSA value (ng/mL) and PSA testing Interval (months), adjusting for Age at First PSA. All Poisson models were adjusted for Agent Orange exposure, service-connected percent, State ADI at first PNBx, race (non-Hispanic Black vs other), and BMI (obese vs not obese) at first PNBx.

**Supplementary Table 6: Sensitivity Analysis Comparison of Unweighted and Inverse Probability of Censoring Weighted Cox Model to Adjust for Selection Bias**

|  | **Unweighted Cox Model aHR (95% CI)** | **Truncated IPCW Cox Model aHR (95% CI)** |
| --- | --- | --- |
| **Age at First PSA**  <50 years  50-59 years  ≥60 years | Ref.  1.40 (1.37 - 1.42)  3.55 (3.48 - 3.62) | Ref.  1.26 (1.23 – 1.28)  3.42 (3.34 – 3.49) |
| **First PSA Value**  ≤1 ng/mL  1.01-2.50 ng/mL  2.51-4 ng/mL  >4 ng/mL | Ref.  1.54 (1.52 – 1.57)  3.31 (3.24 – 3.39)  8.53 (8.35 – 8.72) | Ref.  1.60 (1.57 – 1.64)  3.42 (3.34 – 3.50)  8.82 (8.62 – 9.02) |

Abbreviations: IPCW=Inverse Probability of Censoring Weights, PNBx=Prostate needle biopsy, PSA=Prostate-specific antigen. IPCW were calculated from logistic regression models for the probability of PNBx conditional on covariates (denominator) and conditional on Age at First PSA, First PSA value (numerators). Stabilized weights were truncated at the 1^st^ and 99^th^ percentiles and used to weight Cox models. Both logistic regression models for denominators of stabilized IPCW and Cox models for hazard ratios of metastatic PCa were adjusted for PSA screening interval, Agent Orange exposure, service-connected percent, State ADI at first PNBx, race (non-Hispanic Black vs other), and BMI (obese vs not obese) at first PNBx.
